# Social Vulnerability Does Not Predict 3-Month Functional Outcomes After Mechanical Thrombectomy

**DOI:** 10.64898/2026.01.16.26344311

**Authors:** Nikolas A. Melkumyan, Erick Martinez, Richard Zampolin, Deepak Khatri, Oluwafemi Balogun, Charles Esenwa

## Abstract

**Background:** Mechanical thrombectomy has become standard-of-care in the treatment of emergent large vessel occlusion. However, it is not yet known if social factors impact post-thrombectomy recovery. We studied the association between clinical and sociodemographic factors with 3-month functional outcomes post thrombectomy.

**Methods:** In this prospective cohort study, 291 patients who underwent mechanical thrombectomy at Montefiore-Einstein Hospital in NYC between 1/1/2021 and 4/1/2024 were analyzed. The cohort spanned multiple census tracts across New York City and surrounding areas and included a diverse patient population. The primary outcome was change in modified Rankin Scale (ΔmRS) from pre-stroke baseline to 90–180 days post-stroke. Ordinal logistic regression was used to assess the relationship between ΔmRS and social vulnerability, adjusting for age, sex, stroke severity, and procedural success.

**Results:** Worse functional outcomes were associated with older age (OR 1.03; p = 0.004), male sex (OR 1.85; p = 0.006), higher stroke severity (OR 1.71; p < 0.001), and lower reperfusion success (OR 2.22; p = 0.011). Social vulnerability was not significantly associated with long-term outcomes (OR 0.88; p = 0.550).

**Conclusion:** In this cohort, functional outcomes after mechanical thrombectomy were influenced by clinical and procedural factors rather than sociodemographic vulnerability. While equitable outcomes were observed in the acute setting, ongoing research is needed to explore potential disparities across the broader stroke care continuum, including post-acute recovery.

## Introduction

Stroke remains a leading cause of long-term adult disability with an estimated 795,000 people experiencing a new or recurrent stroke in the United States per year (1). While US stroke mortality has consistently declined in the last 50 years, an aging population is expected to drive stroke incidence up by 2.25 times by 2050 (2). This highlights the continued need for advancements in acute stroke treatment and post-stroke recovery. Perhaps the most significant advancement in stroke care has been the introduction of thrombectomy for emergent large vessel occlusion (ELVO); improving three-month outcomes by 72% compared to standard medical therapy (3). However, mechanical thrombectomy has not been equally accessible to patients from disadvantaged neighborhoods (4, 5, 6); highlighting the importance of community-level factors in stroke disparities.

There is increasing interest in how pre-stroke contextual factors influence recovery after thrombectomy. The Social Vulnerability Index (SVI), developed by the Centers for Disease Control and Prevention (CDC), is a composite measure that incorporates 16 social factors across four domains: socioeconomic status, household composition/disability, minority status/language, and housing/transportation factors (7). SVI has been used in prior stroke research as a proxy for social determinants of health and has been validated for capturing disparities in both access to care and post-stroke outcomes (8,9). Prior studies suggest that higher SVI scores are associated with reduced access to thrombectomy and worse stroke outcomes (5).

The objective of this study was to investigate the influence of demographic, clinical and social variables on long term functional outcomes in patients undergoing mechanical thrombectomy. Leveraging a large and demographically diverse cohort, this study aims to clarify the relative contributions of clinical versus social factors and inform efforts to improve recovery and reduce disparities in stroke care.

## Methods

This prospective cohort study included consecutive patients treated at Montefiore-Einstein Hospital, a Comprehensive Stroke Center, between January 1, 2021 and April 1, 2024. Inclusion criteria were age >18 years with a primary or secondary diagnosis of acute ischemic stroke, confirmed by ICD-10 code, clinical confirmed ELVO treated with mechanical thrombectomy. Mechanical thrombectomy was defined as endovascular clot retrieval in accordance with institutional stroke protocols and irrespective of time windows. Only patients with completed thrombectomy attempts (e.g., documented Thrombolysis in Cerebral Infarction [TICI] scores) and complete outcome measure data for pre-stroke mRS and 90-to-180-day post-stroke mRS were included in the final analysis. Missing data resulted in exclusion from the study cohort, and no data imputation was performed. Out of a potential 322 patients, a total of 291 patients met the inclusion criteria for this study and were included in the final analysis.

Covariates of interest included patient demographics (e.g., age, sex, race/ethnicity), comorbidities, medication use, stroke characteristics, and functional outcome measurements (e.g., pre- and post-stroke mRS) (Supplementary Table 1).

Sociodemographic vulnerability was assessed on the census tract level using the Social Vulnerability Index (SVI), which is derived from U.S. Census data. Each census tract receives a percentile score (0–100), with higher scores indicating greater vulnerability. Patient residential addresses at the time of stroke were linked to their corresponding census tract and SVI score. In our cohort, patients were drawn from census tracts spanning New York City, Westchester County (NY), and New Jersey. A total of 210 census tracts were represented, with SVI values ranging from 5.91 to 99.96 (median: 89.48). Median household income across these census tracts ranged from approximately $11,814 to $250,000+. A full list of variables included in the SVI model, along with their definitions, is provided Supplementary Table 2.

The primary outcome was change in modified Rankin Scale from pre-stroke baseline to 90-180 days post-stroke (ΔmRS), with higher values indicating greater functional decline. This value was treated as an ordinal outcome variable. The cohort was stratified into patients into a low vulnerability and a high vulnerability group using the median SVI score.

Baseline characteristics were summarized using descriptive statistics in Table 1. Continuous variables were reported as means with standard deviations while categorical variables were presented as frequencies and percentages. The cohort had a median SVI score of 89.48 that was used to stratify patients into a low vulnerability (SVI ≤ 89.48) and a high vulnerability (SVI > 89.48) group. In the adjusted model race/ethnicity was removed as an independent variable because of collinearity with social vulnerability. Chi-square was used for categorical variables and either the independent samples t-tests or Wilcoxon rank-sum tests for continuous variables, as appropriate. Corresponding p-values were reported to evaluate statistical significance.

**Table 1:** Baseline Characteristics.

| Baseline Characteristics | Overall (n = 291) | High Vulnerability (n = 145) | Low Vulnerability (n = 146) | p-value |
| --- | --- | --- | --- | --- |
| Age (mean (SD)) | 68.62 (14.77) | 66.50 (15.12) | 70.73 (14.16) | 0.014* |
| Male (%) | 138 (47.4) | 70 (48.3) | 68 (46.6) | 0.863 |
| Race/Ethnicity (%): |  |  |  | <0.001* |
| White or Caucasian | 80 (27.5) | 17 (11.7) | 63 (43.2) |  |
| Black or African American | 105 (36.1) | 57 (39.3) | 48 (32.9) |  |
| Spanish, Hispanic or Latino Origin | 83 (28.5) | 59 (40.7) | 24 (16.4) |  |
| Other | 23 (7.9) | 23 (7.9) | 11 (7.5) |  |
| Previous CVA or TIA | 57 (19.6) | 30 (20.7) | 27 (18.5) | 0.746 |
| Mean Arterial Pressure (MAP) on Admission | 105.66 (19.94) | 106.26 (19.07) | 105.06 (20.81) | 0.608 |
| Past Medical History: |  |  |  |  |
| Hypertension | 225 (77.3) | 108 (74.5) | 117 (80.1) | 0.312 |
| Atrial Fibrillation | 115 (39.5) | 51 (35.2) | 64 (43.8) | 0.164 |
| Dyslipidemia | 111 (38.1) | 44 (30.3) | 67 (45.9) | 0.009 |
| Cardiovascular Disease | 106 (36.4) | 53 (36.6) | 53 (36.3) | 1.000 |
| Left Ventricular Hypertrophy | 68 (23.4) | 33 (22.8) | 35 (24.0) | 0.915 |
| Cardiac Stent Placement | 22 (7.6) | 9 (6.2) | 13 (8.9) | 0.517 |
| Chronic Kidney Disease | 31 (10.7) | 15 (10.3) | 16 (11.0) | 1.000 |
| Antiplatelet Use | 82 (28.2) | 39 (26.9) | 43 (29.5) | 0.723 |
| Admission NIHSS | 16.96 (7.26) | 17.47 (7.65) | 16.45 (6.84) | 0.230 |
| ΔmRS | 2.72 (2.03) | 2.84 (2.10) | 2.60 (1.95) | 0.303 |
| Thrombectomy Characteristics: |  |  |  |  |
| TICI Grade, Unsuccessful (%) | 48 (16.49) | 27 (18.6) | 21 (14.4) | 0.415 |
| Last Known Well to First-Pass Time (mins) | 592.99 (581.21) | 631.33 (650.04) | 555.49 (504.49) | 0.282 |
| Hospital Arrival to Thrombectomy Suite Arrival Time (mins) | 94.89 (141.09) | 99.90 (147.01) | 89.89 (135.24) | 0.550 |
| Hospital Arrival to Thrombectomy Last-Pass/Recanalization Time (mins) | 157.36 (140.69) | 160.40 (144.51) | 154.38 (137.27) | 0.721 |
| Use of Thrombolytics | 101 (34.7) | 52 (35.9) | 49 (33.6) | 0.773 |

\* p < 0.05

All analyses were performed in MyAnalyst (Clinical Reports v1.2.0) and confirmed with R (R version 4.1.1). All assumptions for ordinal regressions were assessed. Outcome variables included odds ratios (ORs) and 95% confidence intervals (CIs) with a two-tailed p-value of < 0.05 being considered statistically significant. The study received Institutional Review Board (IRB) approval from Montefiore Medical Center, and all data was de-identified to protect patient confidentiality.

## Results

We included a total of 291 patients receiving thrombectomy during the study period. Mean age was 68.6 years (SD 14.8). Participants in the high vulnerability group (SVI > 89.48) were younger (mean 66.5 vs. 70.7 years; p = 0.014). Sex distribution was similar between groups (48.3% vs. 46.6% male; p = 0.863). Racial and ethnic differences were significant (p < 0.001): the high vulnerability group had a greater number of Black (39.3% vs. 32.9%) and Hispanic/Latino patients (40.7% vs. 16.4%), while the low vulnerability group had more White patients (43.2% vs. 11.7%). Comorbidities including hypertension, prior stroke/TIA, atrial fibrillation, left ventricular hypertrophy, cardiac stents, chronic kidney disease, and antiplatelet use were not significantly different between the high and low SVI groups. Dyslipidemia was the only comorbidity that showed a statistically significant difference between the two groups where it was more common in the low vulnerability group (45.9% vs. 30.3%; p = 0.009). Thrombectomy-related characteristics such as TICI grade, time from last known well to thrombectomy first-pass, hospital arrival to first pass, and hospital arrival to final pass/recanalization were not statistically significant between the two SVI groups. Other clinical variables such as admission mean arterial pressure (MAP), baseline NIHSS scores, mean ΔmRS, and thrombolytic therapy use also did not differ significantly between groups, as summarized in Table 1.

Ordinal logistic regression demonstrated that increasing age was independently associated with worse functional outcomes, reflected by a shift toward higher ΔmRS scores (OR 1.03; 95% CI: –1.04; p = 0.004) (Table 2). Male sex was also significantly associated with poorer outcomes, with 85% higher odds of worsening ΔmRS compared to females (OR 1.85; 95% CI: 1.19–2.88; p = 0.006). Greater stroke severity, measured by NIHSS, was strongly associated with worse functional outcomes (OR 1.10; 95% CI: 1.06–1.13; p < 0.001). In contrast, successful reperfusion (TICI 2b-3) was the strongest predictor of good outcomes, with unsuccessful reperfusion conferring 2.22-fold higher odds of worse functional outcomes (OR 2.22; 95% CI: 1.20–4.17; p = 0.011). Taken together, these findings highlight age, sex, initial stroke severity, and reperfusion success as independent predictors of functional recovery after thrombectomy.

**Table 2:** Ordinal Logistical Regression for long term outcomes defined by change in MRS.

| Characteristic | OR (95% CI) | p-value |
| --- | --- | --- |
| Age | 1.03 (1.01, 1.04) | 0.004* |
| Sex (Male) | 1.85 (1.19, 2.88) | 0.006* |
| Previous CVA or TIA | 1.04 (0.60, 1.82) | 0.880 |
| TICI Grade** | 2.22 (1.20, 4.17) | 0.011* |
| Time to Thrombectomy † | 1.00 (1.00, 1.00) | 0.921 |
| Admission NIHSS | 1.10 (1.06, 1.13) | < 0.001* |
| Admission Mean Arterial Pressure | 1.00 (0.99, 1.02) | 0.519 |
| Past Medical History: |  |  |
| Hypertension | 1.54 (0.88, 2.71) | 0.133 |
| Atrial Fibrillation | 0.87 (0.52, 1.45) | 0.586 |
| Dyslipidemia | 0.84 (0.52, 1.37) | 0.485 |
| Cardiovascular Disease | 0.76 (0.45, 1.26) | 0.287 |
| Left Ventricular Hypertrophy | 0.79 (0.47, 1.33) | 0.367 |
| Cardiac Stent Placement | 1.20 (0.49, 2.93) | 0.691 |
| Chronic Kidney Disease | 1.15 (0.55, 2.38) | 0.715 |
| Antiplatelet Use | 1.24 (0.74, 2.09) | 0.421 |
| Thrombolytic Use | 0.96 (0.57, 1.60) | 0.862 |
| SVI Score | 0.88 (0.56, 1.35) | 0.550 |

High vulnerability: SVI > 89.48, Low vulnerability: SVI ≤ 89.48
\* p < 0.05

Although there was a trend toward functional outcomes in patients from more vulnerable communities, the SVI score was not statistically significant in predicting recovery after adjusting for all variables of interest (OR 0.88; 95% CI: 0.56–1.35; p = 0.550). A forest plot illustrating the direction and magnitude of key clinical variables and their associations with functional outcomes is shown in Figure 1.

**Figure 1:**
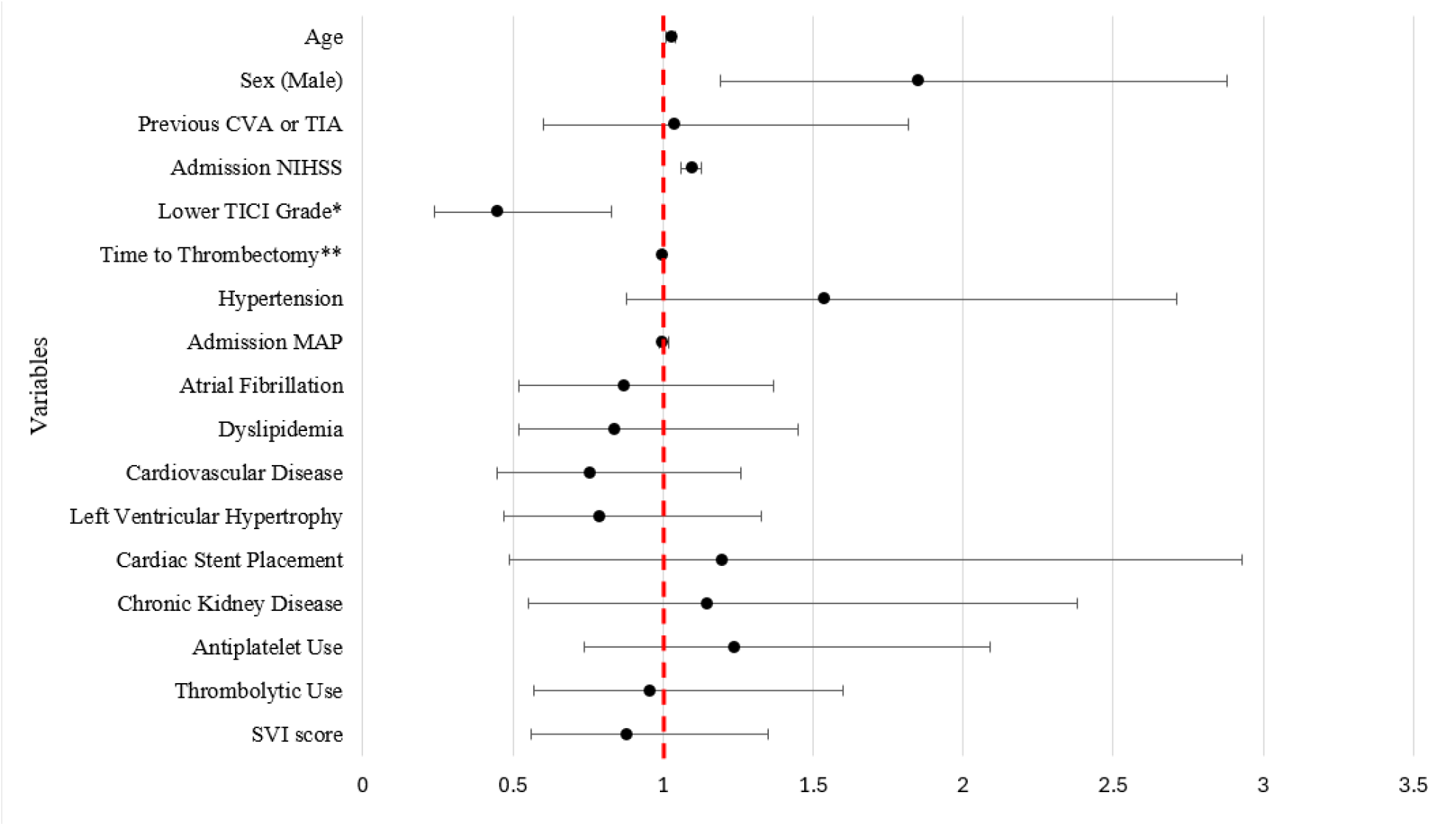
Forest Plot of Ordinal Logistical Regression * Unsuccessful TICI grade defined by TICI 0-2a ** Defined as last known well to time of first pass during thrombectomy procedure

## Discussion

We found that social vulnerability was not associated with 3-month disability outcomes post-thrombectomy in a large diverse single center cohort study. Those with higher social vulnerability were generally younger and more likely to be Black or Hispanic race/ethnicity reflecting broader socio-demographic patterns influencing health vulnerability. While age remained significant as a predictor of long-term outcomes, SVI, a validated marker of community-level vulnerability, was not associated with long term disability post-thrombectomy. While we found significant demographic differences between the dichotomized social vulnerability outcomes, it’s important to note that there was little between-group difference in comorbid conditions (prior stroke, cardiac disease, hypertension), stroke severity (NIHSS), and procedural outcomes (TICI grade). In the fully controlled model older age, male sex, higher NIHSS and lower reperfusion grade were independently associated with poor 3-month outcomes. These findings underscore the importance of pre-hospital and acute stroke care systems in determining stroke outcomes. While social risk is a large driver of stroke risk in the community, our findings suggest that social factors should not be used to influence decision making around offering intervention based on assessed prognostication.

While prior research has highlighted the social and demographic disparities in access to thrombectomy and post-stroke outcomes, our findings suggest that a patient’s functional recovery is more closely linked to stroke severity and procedural success. The absence of a significant association between SVI and functional outcomes also suggests that within this cohort, the clinical decision-making and procedure outcomes for thrombectomies can be considered equitable and not significantly influenced by sociodemographic disparities. Alternatively, the lack of association between SVI and functional outcomes may reflect a measurement limitation in the SVI metric itself. Although stroke severity was similar between the two vulnerability groups, factors such as age and time to presentation (measured through last known well) could have influenced functional outcomes. Examining these variables in future studies may help clarify their role in post-thrombectomy recovery and build upon the findings of this study. Since SVI is based on census tract level data, it could potentially fail to capture individual or household level socioeconomic differences that could potentially play a role in post-stroke recovery. For example, individuals with higher socioeconomic resources who live in higher-vulnerability neighborhoods could have outcomes that differ significantly from their neighbors. Other social factors that could influence patient outcomes and not be detected by SVI include individual education level, healthcare access, income, the type of family support available, and the education level of caregivers. Future studies should incorporate patient-level socioeconomic indicators to better capture the multidimensional impact of social determinants on post-stroke recovery.

Several limitations of this study should be acknowledged. As a single-center, exploratory study, generalizability to regions with different socioeconomic populations is limited. Since the patients were dichotomized into high and low social vulnerability, the stratification may not directly reflect thresholds used in other populations and further limit external generalizability. Additionally, the exclusion of patients due to missing data could mask disparities, especially if this group may have included a higher proportion of socially vulnerable individuals than our study cohort. No analysis of individual SVI subdomains was performed, which may have obscured more specific contributors to social vulnerability Furthermore, no analysis of individual SVI subdomains was performed, which may have obscured more specific drivers of social vulnerability. No formal sample size calculation was performed as part of the statistical analysis and this may reduce the power to detect smaller effects. Outcomes were assessed over a relatively acute timeframe and longer-term follow-up may reveal additional sociodemographic influences. Residual confounding is possible from factors such as cognitive impairment, intensity of post-discharge rehabilitation, and social support. Bias may also arise if socially vulnerable patients were lost to follow-up. Despite these limitations, the lack of observed sociodemographic effects suggests equitable access and procedural performance in thrombectomy-driven care within this cohort. However, disparities may still exist at other stages of care, including stroke recognition, presentation timing, and post-discharge recovery. Future research should incorporate larger, multi-center cohorts and longer follow-up to better assess the long-term impact of social determinants on stroke outcomes and integrating socioeconomic data into acute stroke care may help guide targeted interventions to improve recovery trajectories.

## Data Availability

The data from this study are not publicly available due to IRB restrictions and the presence of potentially identifiable patient information. However, a deidentified version can be made available upon reasonable request.

---

**Supplementary Table 1:**
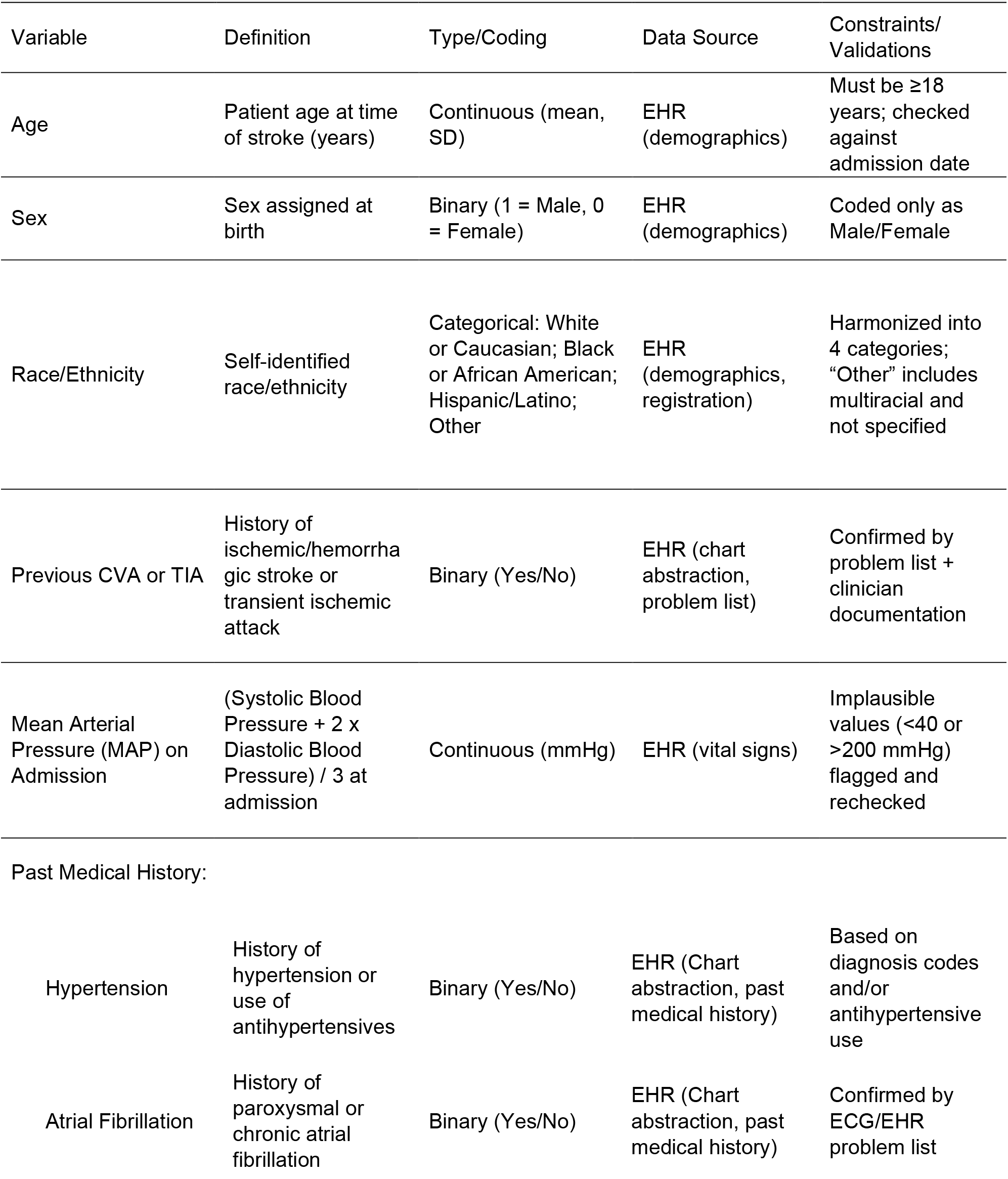
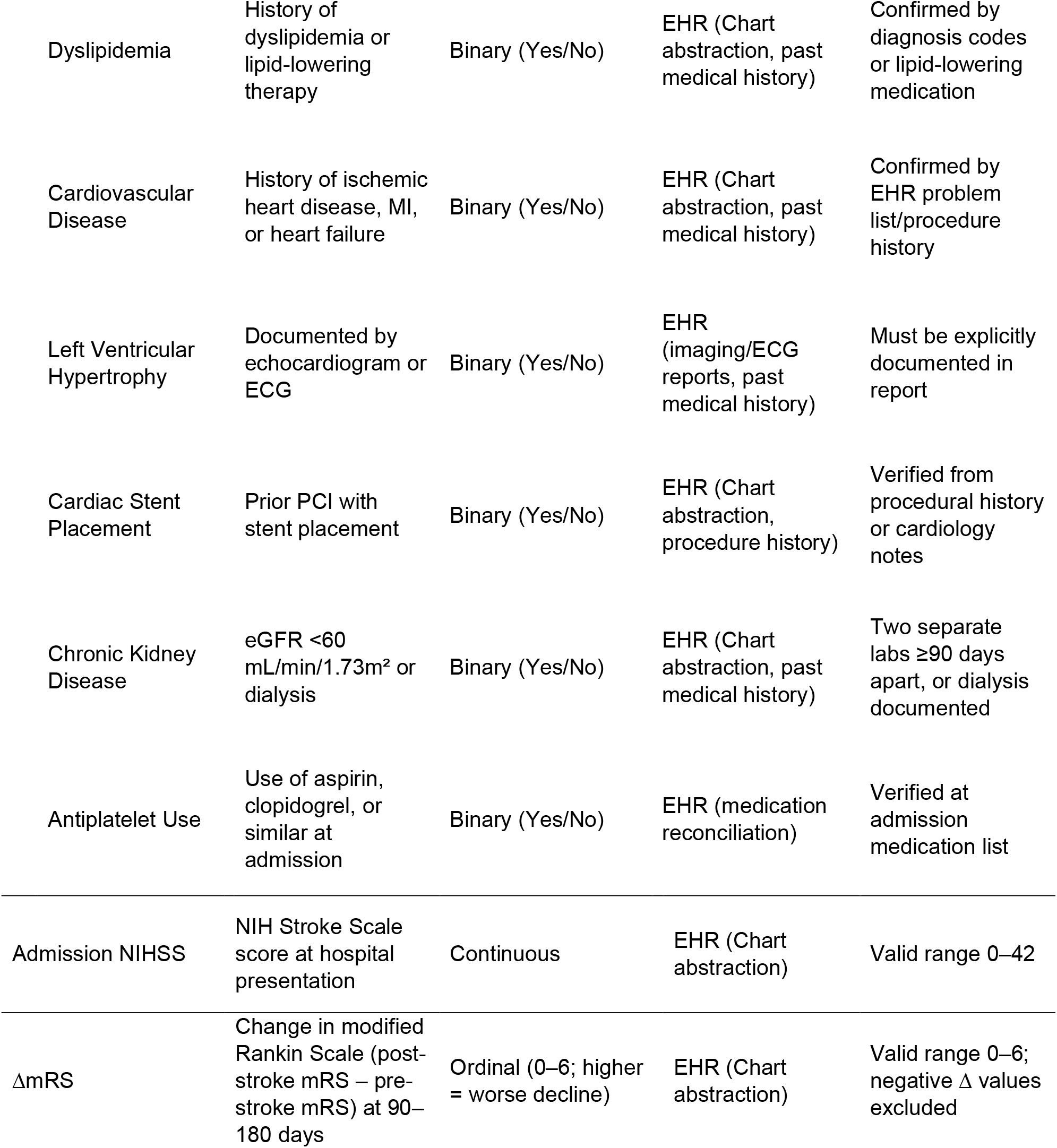
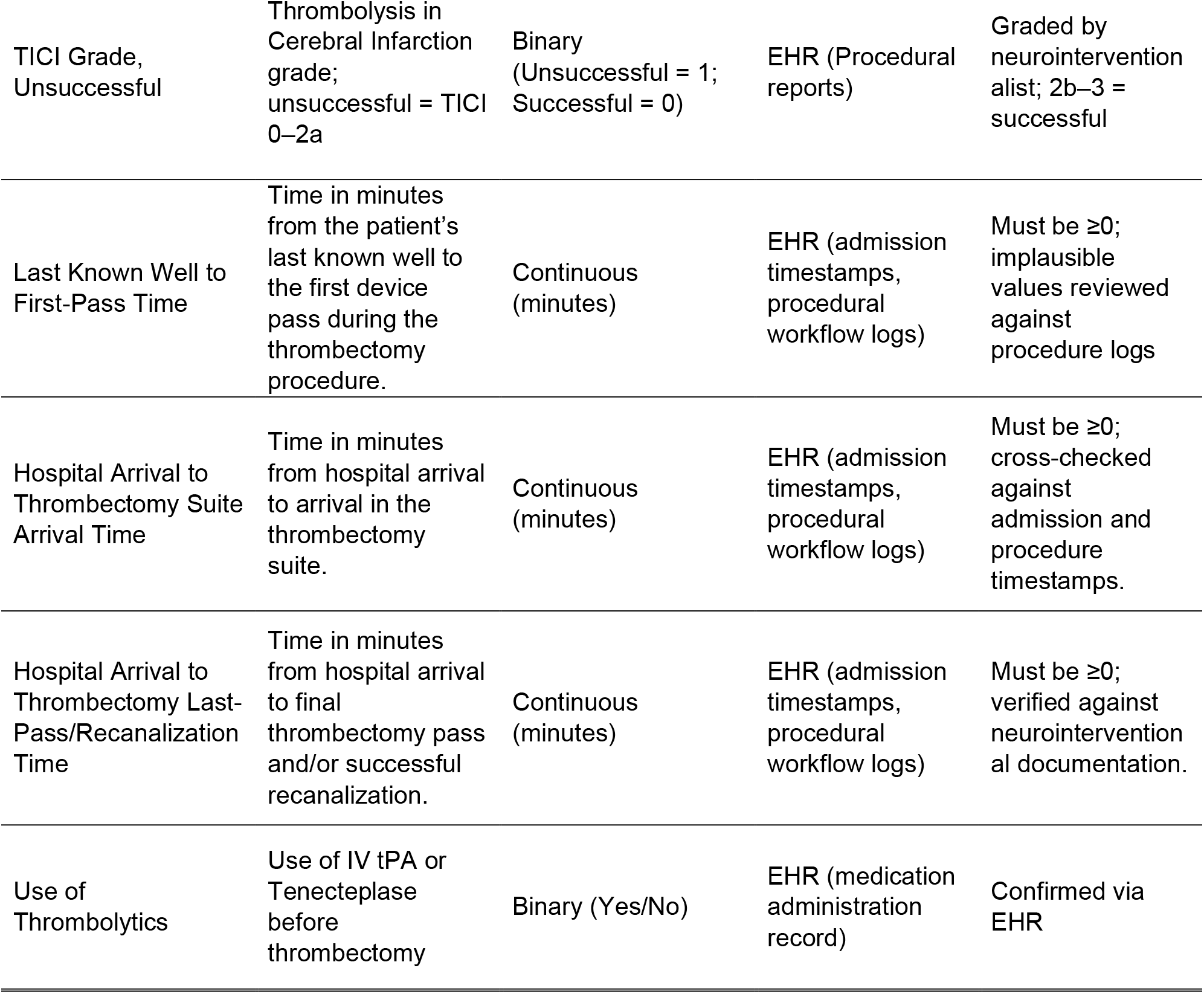
Data Dictionary.

| Variable | Definition | Type/Coding | Data Source | Constraints/<br>Validations |
| --- | --- | --- | --- | --- |
| Age | Patient age at time of stroke (years) | Continuous (mean, SD) | EHR (demographics) | Must be ≥18 years; checked against admission date |
| Sex | Sex assigned at birth | Binary (1 = Male, 0 = Female) | EHR (demographics) | Coded only as Male/Female |
| Race/Ethnicity | Self-identified race/ethnicity | Categorical: White or Caucasian; Black or African American; Hispanic/Latino; Other | EHR (demographics, registration) | Harmonized into 4 categories; “Other” includes multiracial and not specified |
| Previous CVA or TIA | History of ischemic/hemorrhagic stroke or transient ischemic attack | Binary (Yes/No) | EHR (chart abstraction, problem list) | Confirmed by problem list + clinician documentation |
| Mean Arterial Pressure (MAP) on Admission | (Systolic Blood Pressure + 2 x Diastolic Blood Pressure) / 3 at admission | Continuous (mmHg) | EHR (vital signs) | Implausible values (<40 or >200 mmHg) flagged and rechecked |
| **Past Medical History:** |  |  |  |  |
| Hypertension | History of hypertension or use of antihypertensives | Binary (Yes/No) | EHR (Chart abstraction, past medical history) | Based on diagnosis codes and/or antihypertensive use |
| Atrial Fibrillation | History of paroxysmal or chronic atrial fibrillation | Binary (Yes/No) | EHR (Chart abstraction, past medical history) | Confirmed by ECG/EHR problem list |
| Dyslipidemia | History of dyslipidemia or lipid-lowering therapy | Binary (Yes/No) | EHR (Chart abstraction, past medical history) | Confirmed by diagnosis codes or lipid-lowering medication |
| Cardiovascular Disease | History of ischemic heart disease, MI, or heart failure | Binary (Yes/No) | EHR (Chart abstraction, past medical history) | Confirmed by EHR problem list/procedure history |
| Left Ventricular Hypertrophy | Documented by echocardiogram or ECG | Binary (Yes/No) | EHR (imaging/ECG reports, past medical history) | Must be explicitly documented in report |
| Cardiac Stent Placement | Prior PCI with stent placement | Binary (Yes/No) | EHR (Chart abstraction, procedure history) | Verified from procedural history or cardiology notes |
| Chronic Kidney Disease | eGFR <60 mL/min/1.73m<sup>2</sup> or dialysis | Binary (Yes/No) | EHR (Chart abstraction, past medical history) | Two separate labs ≥90 days apart, or dialysis documented |
| Antiplatelet Use | Use of aspirin, clopidogrel, or similar at admission | Binary (Yes/No) | EHR (medication reconciliation) | Verified at admission medication list |
| Admission NIHSS | NIH Stroke Scale score at hospital presentation | Continuous | EHR (Chart abstraction) | Valid range 0–42 |
| ΔmRS | Change in modified Rankin Scale (post-stroke mRS – pre-stroke mRS) at 90–180 days | Ordinal (0–6; higher = worse decline) | EHR (Chart abstraction) | Valid range 0–6; negative Δ values excluded |
| TICI Grade, Unsuccessful | Thrombolysis in Cerebral Infarction grade; unsuccessful = TICI 0–2a | Binary (Unsuccessful = 1; Successful = 0) | EHR (Procedural reports) | Graded by neurointerventionalist; 2b–3 = successful |
| Last Known Well to First-Pass Time | Time in minutes from the patient's last known well to the first device pass during the thrombectomy procedure. | Continuous (minutes) | EHR (admission timestamps, procedural workflow logs) | Must be ≥0; implausible values reviewed against procedure logs |
| Hospital Arrival to Thrombectomy Suite Arrival Time | Time in minutes from hospital arrival to arrival in the thrombectomy suite. | Continuous (minutes) | EHR (admission timestamps, procedural workflow logs) | Must be ≥0; cross-checked against admission and procedure timestamps. |
| Hospital Arrival to Thrombectomy Last-Pass/Recanalization Time | Time in minutes from hospital arrival to final thrombectomy pass and/or successful recanalization. | Continuous (minutes) | EHR (admission timestamps, procedural workflow logs) | Must be ≥0; verified against neurointerventional documentation. |
| Use of Thrombolytics | Use of IV tPA or Tenecteplase before thrombectomy | Binary (Yes/No) | EHR (medication administration record) | Confirmed via EHR |

**Supplementary Table 2:** Components of Social Vulnerability Index (SVI)

| Domain | Variables included | Description |
| --- | --- | --- |
| Socioeconomic Status | Below 150% poverty<br>Unemployed<br>Housing cost burden<br>No high school diploma<br>No health insurance | Captures financial hardship, educational attainment, and access to health coverage. |
| Household Characteristics | Aged 65 and over<br>Aged 17 and younger<br>Civilian with a disability<br>Single-parent households<br>Limited English proficiency | Reflects age- and disability-related vulnerability and household dependency. |
| Racial and Ethnic Minority Status | Hispanic or Latino (any race)<br>Black or African American*<br>Asian*<br>American Indian/Alaska Native*<br>Native Hawaiian/Pacific Islander*<br>Two or more races*<br>Other races, not Hispanic/Latino | Captures minority status and language barriers that may affect access to care. |
| Housing Type and Transportation | Multi-unit structures<br>Mobile homes<br>Crowding (>1 person/room)<br>No vehicle available<br>Group quarters | Measures housing stability, density, and access to transportation. |

\* Non-Hispanic or Latino

